# Providing a safe, in-person, residential college experience during the COVID-19 pandemic

**DOI:** 10.1101/2021.03.02.21252746

**Authors:** Scott A. Travis, Aaron A. Best, Kristyn S. Bochniak, Nicole D. Dunteman, Jennifer Fellinger, Peter D. Folkert, Timothy Koberna, Benjamin G. Kopek, Brent P. Krueger, Jeff Pestun, Michael J. Pikaart, Cindy Sabo, Alex J. Schuitema

## Abstract

Due to the COVID-19 pandemic, higher education institutions were forced to make difficult decisions regarding the 2020-2021 academic year. Many institutions decided to have courses in an online remote format, others decided to attempt an in-person experience, while still others took a hybrid approach. Hope College (Holland, MI) decided that an in-person semester would be safer and more equitable for students. To achieve this at a residential college required broad collaboration across multiple stakeholders. Here, we share lessons learned and detail Hope College’s model, including wastewater surveillance, comprehensive testing, contact tracing and isolation procedures, that allowed us to deliver on our commitment of an in-person, residential college experience.

## Introduction

In the middle of the COVID-19 pandemic, and months before the start of the fall 2020 semester, Hope College President, Matthew Scogin, committed to students and families to do everything possible “to provide an in-person experience for all our students, which includes in-person classes and on-campus living” [1]. On May 20, 2020, a framework was shared to re-open the campus for safe, in-person living and learning. It was our intent that this framework would lead to a safer and more equitable learning environment for all students. A recent Gallup study across higher education suggests it may also be a better one, finding that students who transitioned from an in-person learning environment to online learning said the quality of their education experience declined [2]. Anecdotally, but in agreement with the Gallup study, Hope College professors reported that when the college was online during the Spring 2020 semester students could be seen taking exams in cars outside the local library due to a lack of reliable internet access at home.

Our framework for a return to an in-person college experience for the 2020-2021 academic year included starting classes two weeks earlier than normal and reducing break days to complete the semester before Thanksgiving, adapting instructional spaces, implementing safeguards and health screens with accountability, and frequently communicating with students, families, and employees. Our plans also included strategies for student testing, contact tracing, and isolation. The outcomes and lessons learned from these strategies are outlined here with the hope that others can learn from our work to provide safe, in-person learning experiences of their own (Figure 1).

**Figure 1.**
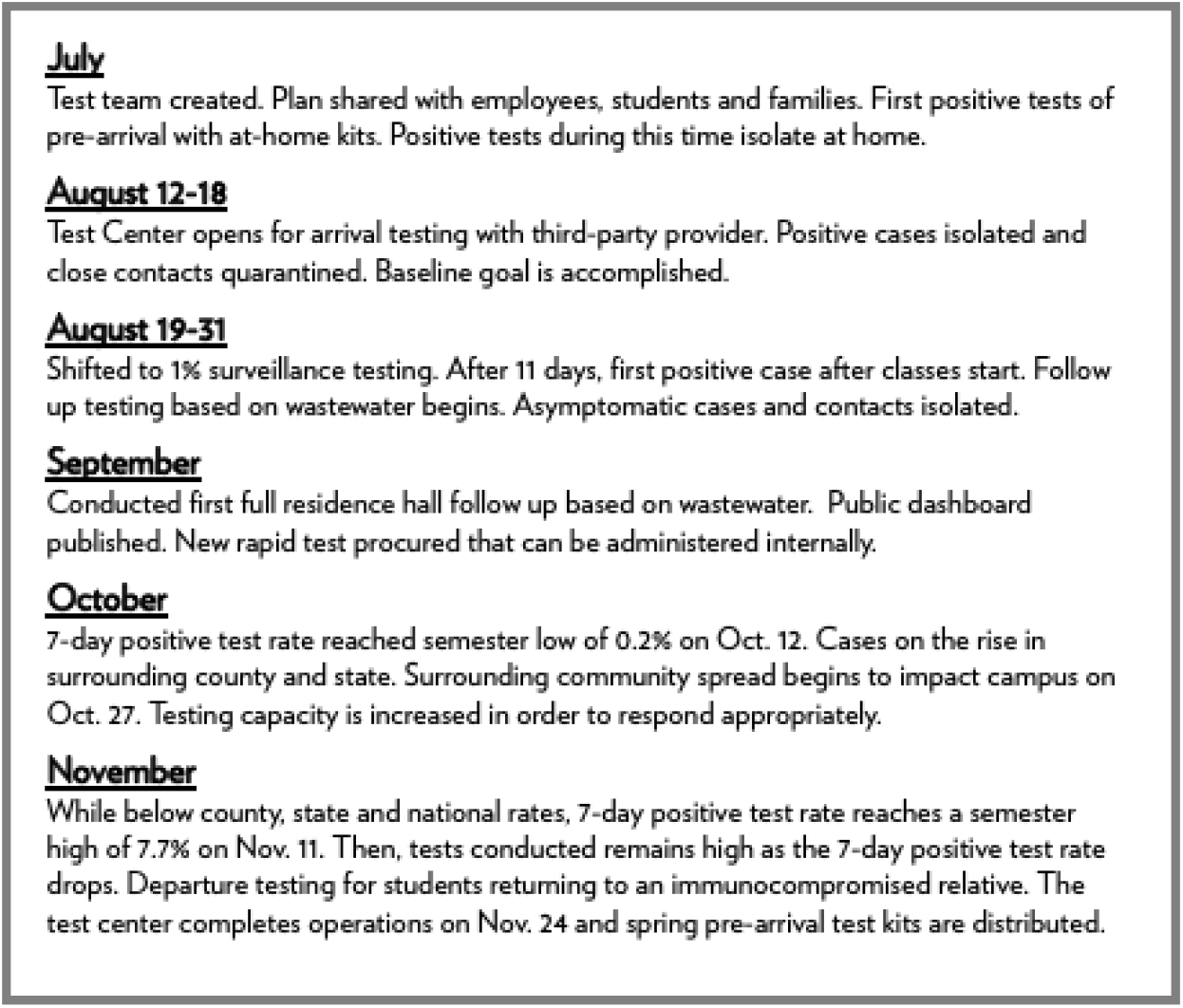
Timeline overview of Hope College’s Fall 2020 semester highlighting various aspects of our mitigation strategy and major events.

### Comprehensive Team Approach

As we prepared for the fall semester, it became clear that we would need to work together across multiple areas of the organization if our framework were to succeed. In July, a team was formed that consisted of two biology faculty members, two IT staff members, a residential life staff member, the head athletic trainer responsible for contact tracing, a vice president responsible for COVID-19 response and public affairs, the director (registered nurse) of the campus health center, and a team lead assigned from another area of college administration. Meeting at least three times a week throughout the semester, this group made sure that the testing, contact tracing, and quarantine and isolation aspects of our framework acted as a single process. Having a diverse cross-functional team played a critical role in sharing and interpreting data from multiple sources in order to take decisive actions when needed.

### Testing

Between July 29 and November 24, 2020, we conducted 10,700 tests at no cost to students and employees. Our testing plan was tailored for our community and informed by the expertise of public health officials and our faculty and staff. We recognized that challenges could arise in both the supply chain for testing equipment and the turnaround time for test results. For this reason, it was critical to take a multi-faceted approach. Our testing plan was meant to supplement and monitor, not replace, all of the other protections put in place as part of our broad pandemic mitigation strategy.

### Baseline Testing

Our first goal was to start the academic year with zero cases of COVID-19 on campus. To achieve a baseline of zero, students and employees were tested with an at-home kit sent directly to them [3] The kit featured a saliva-based test that was medically-supervised via Zoom. Partnering with Vault Health (NY), over 3,000 students and employees were tested 8-10 days before arrival. Because this test captured the result for only that particular moment in time, students and employees were expected to do everything possible to minimize their risk of exposure to the virus as they prepared to.arrive on campus.

Results of the tests were communicated only to the student and members of the test team in accordance with HIPAA guidelines. The housing office was provided a list that indicated if a student was cleared to move in or needed testing upon arrival. Those that arrived on campus without a test result were required to test at a newly opened Test Center adjacent to campus and staffed with a third party contract and administrative college staff reassigned from other areas.

Between the at-home and on-arrival testing, a total of 3,878 tests were administered as part of baseline testing with 38 positive cases identified and isolated either at home or immediately upon arrival. On the first day of classes (August 17, 2020), our 0.98% positive test rate was lower than the national positive rate of 6.1% and state positive rate of 2.5% according to the Johns Hopkins Coronavirus Resource Center [4] (Figure 2).

**Figure 2.**
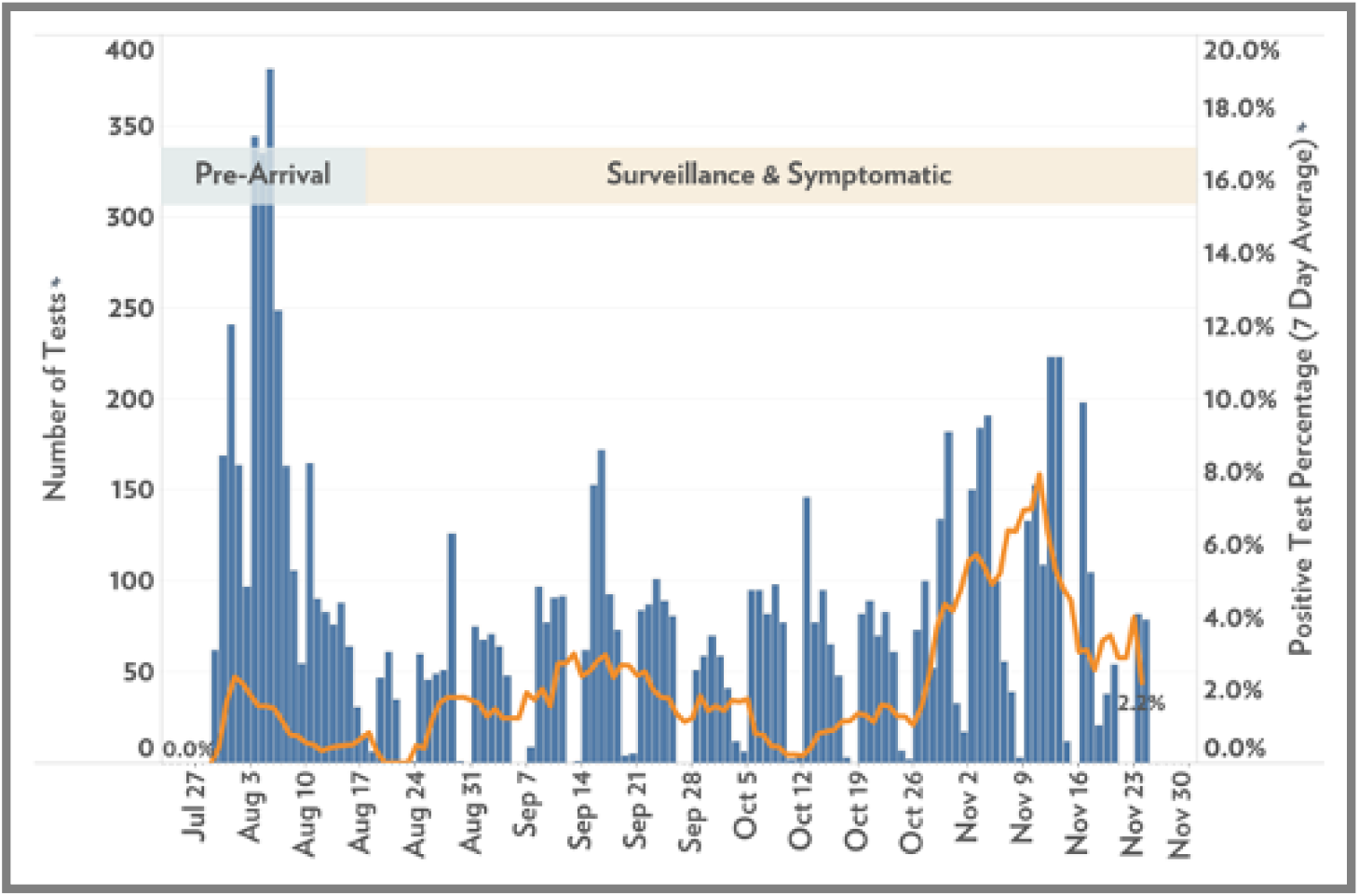
Plot displaying the number of tests conducted and positive-test percentage (7-day average) at Hope College (MI) before and during the Fall 2020 semester.

### Surveillance (asymptomatic) Screening

Our surveillance testing plan was developed to monitor the campus for the presence of SARS-CoV2, catching as many asymptomatic cases as possible, and containing the spread of the virus. As testing strategies were being developed, several mathematical models were published to predict the effectiveness of viral transmission mitigation strategies [5,6]. In these models, the greatest reduction in case number was achieved by many of our strategies (i.e., mask wearing, physical distancing, etc.). Therefore, we determined that a lower rate (1% of campus population daily) of surveillance screening would be sufficient to detect outbreaks in campus housing. This is a much lower testing frequency than advocated in some other models [7], but was determined to be our best strategy based on our constraints. We were able to contract with a service provider to enable rapid (15 min) testing for surveillance. This testing service allowed for a maximum of 60 tests to be run per day. As new testing methods were introduced and reagents produced nationwide, we were able to begin performing rapid (15 minute) antigen tests. Rapid antigen tests were run by the Hope College Health Center nursing team. When we felt they were needed, we were able to perform large-scale testing events. Early in the semester, saliva-based PCR testing was used to conduct one of these testing events. As the semester progressed, testing events used the rapid antigen tests.

For surveillance testing, students were selected at random and the testing took place at the campus Test Center every weekday. Notifications and reminders were sent from a dedicated Test Center email account to the student’s college-provided email account. In most cases, students had up to 48 hours to complete their test.

### Wastewater Surveillance Testing

Between August 27 and September 6, the wastewater surveillance testing program was ramped up with coverage of approximately 55% of the entire student population, including 70% of those in college-owned housing. This led us to switch to a more targeted testing strategy based on wastewater data for the covered student population. Those living outside of the wastewater testing zones continued to be screened through randomized surveillance testing.

Wastewater coming from nine specific residential zones on campus was collected using dedicated autosamplers each weekday, yielding 24 hour composite samples. Each zone had between 100 to250 residents. Wastewater samples were collected and analyzed for the presence of viral genomic material by quantitative PCR. This method allowed for same-day results regarding the presence or absence of viral genetic material in the campus wastewater zones. Thus, we were able to react quickly in response to infected individuals within campus housing even if those individuals were not showing symptoms. Follow-up testing of individuals based on wastewater samples was conducted on 29 different occasions between August 31 and November 16. On multiple occasions, as testing capacity allowed, entire residential halls were asked to test within 24 hours.

Including both the 1% sample and individual wastewater follow up testing, 5,696 surveillance tests were conducted during the semester, resulting in 57 positive cases (a 1% positive rate) (Figure 2). It is important to note that these were asymptomatic cases identified and isolated, with contact tracing leading to additional quarantined students, who were unable to infect others.

### Subset Testing

There were some groups of individuals, including residential life staff and student-athletes, that were tested regularly. While the Michigan Intercollegiate Athletic Association postponed conference competition, our athletic teams continued team activities and conducted additional testing regularly. This was completed by athletic training staff in partnership with the overall testing process. An additional testing subset at the end of the semester included students who were returning to a home where there was an immunocompromised family member.

### Symptomatic Testing

Symptomatic testing was reserved for students who experienced and reported symptoms of COVID-19. These tests were conducted by registered nurses from the Hope College Health Center. Students were asked to self-quarantine until they received a test and result. Employees experiencing symptoms were asked to test through their healthcare provider. During the semester, 960 symptomatic tests were conducted, resulting in 124 positive cases. The positive rate for symptomatic tests was 12.9%.

### Screening Form

Students who would be on campus were asked to complete a screening form that asked if they were experiencing any COVID-like symptoms. Students were asked, but not required, to complete this form. Participation in this screening form dropped from 1882 submissions on the first day of classes to 316 on the final day of classes. Employees who were going to be on campus were required to complete a daily screening form for COVID-19 symptoms as directed by the Michigan Department of Occupational Health and Safety. If students were experiencing symptoms they were directed to the Campus Health Center while employees were directed to see their physician.

### Contact Tracing

Students were also required to participate in the contact tracing process. Each positive case prompted a contact tracing investigation to determine close contacts. Close contacts were defined as individuals who had been within 6 feet of a positive case for a cumulative total of 15 minutes or more. As we began the semester, trained advocates, often college staff from other areas, helped students begin the process of identifying close contacts so that they were prepared to work with the health department. While the Ottawa County Department of Public Health remained a close partner all semester long, these investigations shifted increasingly to college staff as contact tracing resources in the surrounding community became unavailable.

This contact tracing played a critical role in mitigating the spread of the virus and our ability to conduct our own investigations throughout the semester was a critical component that allowed us to continue in-person. A contact tracing team of 7 individuals conducted over 150 investigations throughout the semester leading to 670 close contacts in quarantine. While our semester average of close contacts per positive case was between 4 and 5, we identified a trend of more close contacts per positive case in the latter part of the semester as, presumably, student adherence to safeguards lessened.

### Isolation and Quarantine

All positive cases and close contacts were isolated or quarantined to limit the spread of COVID-19 on campus. Isolation separated infected individuals from others and lasted 10 days from the first date of symptoms (or test date for asymptomatic cases). Quarantine separated and restricted the movement of people who were close-contacts with a known infected individual and lasted 14 days from last known contact. In general, isolation and quarantine resulted in students leaving their campus housing and moving into designated housing. However, in some cases (apartments, cottages) where all roommates were considered close contacts students had the option to remain in their original housing. Students were also allowed to go home to their permanent residence to quarantine or isolate unless instructed otherwise by the local health department. At the start of the semester, the college reserved 126 rooms for isolation and quarantine purposes. This increased to 176 rooms by the end of the semester. The peak of students in isolation and quarantine, including those in isolation or quarantine at home, was 369 on November 11.

A team of trained advocates and healthcare professionals supported students who were in isolation or quarantine to make their experience as comfortable as possible. These advocates helped with moving, informed students of resources, checked in on them, facilitated communication with faculty, and helped them understand their role in the contact tracing process. While in isolation or quarantine, students participated in classes remotely. Faculty were prepared to engage students in their courses using online tools. Dining services created a special menu and delivery service. Symptomatic students were asked to take their temperature and asymptomatic students were asked to monitor for symptoms. The process, including the availability of housing and advocates, applied to all students, whether they resided on or off-campus.

We used a symptom-based and time-based strategy, not a test-based strategy, to determine a return date for individuals diagnosed with confirmed or suspected COVID-19. This strategy took into account the time since the diagnosis and the time since recovery as well as the presence or absence of symptoms. The decision to end isolation or quarantine and return to campus was made in consultation with healthcare providers and the local health department.

We observed 21 cases where a close contact student that went into isolation became symptomatic and tested positive. Thus, quarantining close contacts likely reduced the number of infections on campus. Our experience serves an example of how contact tracing and quarantine procedures worked to mitigate spread.

## Discussion

As we prepared to continue in-person learning during the spring of 2021, we considered the following lessons and offer them to others preparing for similar situations:

- Information must be available and actionable. The teams involved in the various stages of the process (from wastewater results to quarantine capacity) need a consistent and accurate way to share information, interpret available data, and make data-driven decisions.
- Diverse perspectives lead to better decision-making. Having representatives from each area regularly meet to share information, have honest and difficult discussions, and make recommendations to decision-makers helps make sure we make the right decisions at the right time.
- A good working relationship with the local health department is critical. We follow the recommendation of the Centers for Disease Control and Prevention to work with our local health department and are fortunate to partner with the Ottawa County Department of Public Health. Meetings and follow up conversations are frequent and comprehensive.
- Constraints must be acknowledged and managed. We try to start with the ideal approach and work backward based on identified constraints. These could include testing capacity, staffing, housing capacity, regulations, or finances.
- Talented teams make difficult work possible. Including the COVID-19 Steering Committee and sub-teams around wastewater, testing operations, contact tracing, academics, safety operations, and housing, it is estimated that 150 employees, approximately 20% of our workforce, have had at least part of their job realigned to respond to COVID-19.

During a meeting on October 30, amid a local, state, and national outbreak, local health officials confirmed that our students were safer on campus within our framework than they were elsewhere. The Ottawa County Department of Public Health shared that while our campus situation reflected the reality of the broader West Michigan region where viral spread was picking up rapidly, because it is a highly controlled environment Hope was actually better positioned than our surrounding communities. We were able to quickly identify areas of viral spread, schedule tests, isolate and quarantine individuals, complete contact tracing, and notify close contacts very effectively and efficiently. This stands in contrast to what may have been experienced in communities with large universities (>20,000 students) where the incidence of infection was not sufficiently contained [8]. Therefore, we recommend that all higher education institutions seek to implement a comprehensive framework similar to the one outlined here and implemented at Hope College.

## Data Availability

Data is available upon request.

## Acknowledgments

Funding for some wastewater analysis was provided by the Michigan Department of Environment, Great Lakes and Energy. The funders had no role in study design, data collection and analysis, decision to publish, or preparation of the manuscript.

## Notes

### Competing Interest Statement

The authors have declared no competing interest.

### Funding Statement

Funding for wastewater testing was provided by the Michigan Department of Environment, Great Lakes and Energy

### Author Declarations

The Hope College Institutional Review Board approved this analysis.

